# Meconium androgens are correlated with ASD-related phenotypic traits in early childhood

**DOI:** 10.1101/2020.03.05.20031609

**Authors:** Dina Terloyeva, Alexander J. Frey, Bo Y. Park, Elizabeth M. Kauffman, Leny Mathew, Anna Bostwick, Erika L. Varner, Brian K. Lee, Lisa A. Croen, Margaret D. Fallin, Irva Hertz-Picciotto, Craig J. Newschaffer, Kristen Lyall, Nathaniel W. Snyder

## Abstract

Prenatal exposure to increased androgens has been suggested as a risk factor for autism spectrum disorder (ASD). This hypothesis has been examined by measurement of steroids in amniotic fluid and cord blood, with mixed results. To provide an orthogonal measure of fetal exposure, this study used meconium, the first stool of a newborn, to measure prenatal androgen exposure from infants in the Early Autism Risk Longitudinal Investigation (EARLI). EARLI is a familial-enriched risk cohort that enrolled pregnant mothers who already had a child with an ASD diagnosis. In the younger child, we investigated the association between meconium unconjugated (u) and total (t) concentrations of major androgens testosterone (T), dehydroepiandrosterone (DHEA) and androstenedione (A4), and ASD-related traits at 12 and 36 months of age. Autism traits were measured at 12 months with Autism Observation Scale for Infants (AOSI) and at 36 months with total score on the Social Responsiveness Scale (SRS). 137 children (61 males, 76 females) had data on both outcomes and meconium androgen measurements. Separate robust linear regressions between each of the log-transformed androgens and log-transformed AOSI and SRS scores revealed three-way interaction between sex of the child, sex of the proband, and androgen concentration. In the adjusted analyses; t-T, u-A4, and u-DHEA (*P* ≤ 0.01) were positively associated with AOSI scores while u-T (*P*=0.004) and u-DHEA (*P*=0.007) were positively associated with SRS total score among females with female probands. Additionally, higher concentrations of u-T (*P*=0.01) and t-T (*P*=0.01) predicted higher SRS total score in males with male probands.

**Significance:** Using a prospective pregnancy cohort enriched for autism risk, we investigated prenatal androgen exposure measured from meconium as a risk factor for autism-related traits. Several meconium androgens were positively correlated with autism-related traits. In addition, we found a strong positive association between autism traits in the sub-group of individuals with an older female sibling with autism extending a previous finding based on cord blood measures in the same cohort. This study supports the utility of meconium for studies of endogenous fetal metabolism and suggests the sex of the proband should be considered as a biological variable in relevant studies.

## Introduction

Autism spectrum disorder (ASD) has an estimated prevalence of 1 in 59 among 8-year old children in the United States.(1, 2) Within this population level estimate, there is a strong sex-dependent bias with a 4:1 male:female ratio.(1, 3, 4) The mechanism underlying this sex difference remains unknown but is of interest in understanding the etiology of ASD. Critically, it is unknown whether female-specific protective factors, male-specific risk factors, or both drive this difference. The strong male sex bias in the prevalence of ASD gave rise to two theories: “extreme male brain” theory and multiple threshold liability model.

Some researchers argue that sex differences between ASD-affected children mirror differences between males and females in the general population. Combined with evidence that ASD-affected children outperform typically developing children at a range of intellectual tests,(5, 6) these observations form one tenet of the extreme male brain theory. According to this theory, a “male brain” type is better at spatial tasks than social ones and at systematizing rather than empathizing, and an autistic brain is an extreme type of this “male brain”.(7)

The multiple threshold liability model suggests that females are protected from exhibiting autistic traits severe enough to qualify for an ASD diagnosis, even with an accumulated etiological load sufficient for males to reach a diagnostic threshold.(8, 9) Female-specific protective factor(s) have not yet been established; however, two findings support this hypothesis. Firstly, it has been shown that ASD-affected females carry a greater load of de novo copy-number mutations compared to ASD-affected males.(10) Secondly, siblings of affected females demonstrate higher autistic behavior scores compared to siblings of affected males.(11-13)

The strong sex differential in ASD prevalence also suggests that developmental processes of early sex-differentiation interact with etiological events driving developmental changes of ASD that manifest later in life. Indeed, multiple studies implicate the fetal development stage in the origin of this disorder and link genetic influences, neuroanatomical changes, and environmental influences such as steroid exposures to ASD outcome.(14-21) Steroid hormones are crucial for sex-differentiation of the human fetus and development of the central nervous system; therefore, several studies have suggested a potential link between abnormal steroid hormone levels and ASD. Androgens, the male sex steroids that bind to the androgen receptor, have received the most study.

Previous studies measured androgen levels in either amniotic fluid or cord blood to estimate fetal exposure to the male sex steroids. In two separate studies, the same group found a positive correlation between amniotic fluid testosterone levels and autism assessment scales.(22, 23). Other studies using either amniotic fluid or cord blood reported mixed, but mostly null results. (24-27)

To overcome the limitations of amniocentesis and cord blood, we used meconium, the first stool of a newborn, to estimate prenatal androgen exposure. Since formation of meconium is thought to start around the 12-13^th^ week of gestation, and meconium can be non-invasively collected within 72 hours of the birth of the child, meconium has the potential to capture cumulative prenatal exposure.(28). Previous studies have utilized meconium samples to examine prenatal exposure to both exogenous and endogenous compounds,(29-31) and we recently developed a robust method to quantify androgens in human meconium.(28)

This study investigated the association between levels of androgens, namely, testosterone (T) and its weakly androgenic precursors, dehydroepiandrosterone (DHEA) and androstenedione (A4), measured in meconium, and autistic traits at 12 and 36 months of age. Since these steroids exist in both unconjugated and glucuronide and/or sulfate-conjugated forms, and previous research has suggested glucuronidation may be different in autistic individuals,(32) we performed analysis on both unconjugated (u) and total (t) forms that include both conjugated and unconjugated. Our study sample was drawn from the Early Autism Risk Longitudinal Investigation (EARLI), an enriched ASD risk pregnancy cohort where enrolled families had an older child (the study proband) with an ASD diagnosis. Park et al. also analyzed samples from the EARLI cohort and did not find an association between prenatal androgens measured in cord blood and ASD phenotype; however, stratification by the proband’s sex revealed a positive relationship between testosterone levels and ASD phenotype among participants whose older, affected sibling was a female. (27) Based on this previous sub-group analysis we examined effect modification by sex of the child and sex of the proband.

## Methods

### Study population

This study used data and biosamples from the Early Autism Risk Longitudinal Investigation (EARLI). EARLI design and study population have been reported elsewhere.(33) In short, the study included mothers of children with ASD diagnoses: autistic disorder, Asperger syndrome, or pervasive developmental disorder not otherwise specified, who were pregnant at the time of recruitment. Pregnant women were recruited by 28^th^ week of pregnancy and followed closely until their child reached 3 years of age. In addition to having a biological child with ASD diagnosis, and being no more than 28 weeks pregnant, women had to be of at least 18 years of age, able to speak English or Spanish, and live within 2 hours of a study site. Recruitment was performed by four participating study centers: Drexel/Children’s Hospital of Philadelphia, John’s Hopkins/Kennedy Krieger Institute, UC Davis; and Northern California Kaiser Permanente, which represented Southeast Pennsylvania, Northeast Maryland, and Northern California.

The study staff provided mothers with meconium collection kits prior to delivery and made arrangements with birth delivery staff at the hospitals and birth centers to assist with sample collection and temporary storage. Meconium was collected as soon as it was passed and was stored in the hospital/birth center or at home in the freezer until it was retrieved by the study staff. The study enrolled a total of 236 mothers with 8 mothers having twins, which resulted in 244 children in total. One of the twins in each twin pair was removed from the dataset at random resulting in 236 children. Of these 236 children, 62 lacked meconium, 29 lacked AOSI assessment at 12 months, and 65 lacked SRS assessment at 36 months resulting in analytic sample size of 137 participants (61 females, 76 males).

### Laboratory methods

Meconium androgens were quantified by liquid chromatography-high resolution mass spectrometry (LC-MS/HRMS) as previously described.(28) Briefly, both unconjugated (u) and total (t) (unconjugated plus conjugated) androgen levels were measured. T, A4, DHEA and internal standards (^13^ C3-T, ^13^ C3-A4, and ^2^ H5-DHEA) were extracted from approximately 50 mg of each meconium sample by methanol, hexane:dicholoromethane (3:2), followed by derivatization with Girard P reagent and LC-MS/HRMS analysis as previously described. Limit of Quantification (LOQ) was defined as 10 times the lowest point on the calibration curve. LOQ was 2.4 ng/g for u-T, t-T, u-A4, and t-A4, and no samples fell below LOQ for DHEA (12 ng/g). Measurements of t-T fell below the limit of quantification defined at 2.4 ng/g for 7 participants (3 boys and 4 girls). For these participants we used values of t-T replaced by 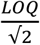

### Outcome assessment

Outcomes included autistic traits measured at 12 months with the Autism Observation Scale for Infants (AOSI) (34) and at 36 months with the caregiver reported Social Responsiveness Scale (SRS), pre-school version.(35, 36) AOSI is a semi-structured direct examiner-administered observational measure intended to capture early signs of autism in children aged 6-18 months. AOSI contains 18 items representing domains of behavior such as emotional responses, eye contact, visual tracking, social interest, motor and sensory behavior. Of these 18 items, 14 are assigned the score from 0 to 3, where 0 represents typical behavior, and 3 represents a lack of behavior. The other four items, which include assessment of eye contact and atypical motor and sensory behavior, are rated on a scale of 0 to 2. Total score on the AOSI scale demonstrated high interrater reliability (unweighted kappa = 0.93) and acceptable test-retest reliability (intra-class correlation = 0.61) at 12 months with a score of 9 and greater was reported to be predictive of ASD diagnosis at 36 months (34).

SRS is a 65-item questionnaire completed by a caregiver at 36 months of age. Items on the SRS mainly assess reciprocal social behavior (RCB), and whether restricted or stereotypical behaviors and communicative deficits impact the child’s ability for RCB.(37, 38) Each item on the scale is assigned a score from 0 to 3, where 0 represents “never true”, and 3 represents “almost always true”. The resulting total raw score ranges from 0 to 180 and higher scores indicate more severe social deficit. SRS has good psychometric properties, and performs well among the general population of children and in children with older ASD-affected siblings.(37-39) Validation of the scale against the Autism Diagnostic Interview-Revised (ADI-R) showed high correlation between SRS scores and ADI-R scores.(37)

### Covariates

Variables previously associated with ASD-phenotype were assessed for potential confounding: maternal age, race, ethnicity, education, income, gestational age, interpregnancy interval, number of previous pregnancies, and caesarian delivery. Assessment of covariates was performed in two steps. At first step, tests for independence were performed between each of the covariates and both exposures and outcomes before the decision was made to include them into the multivariable model. If both exposures and each of the outcomes were different between different levels of covariates irrespective of the p-value, they were carried on to the second step. If the covariate was continuously measured, the correlation coefficient with both the exposure and the outcome had to be greater than 0.2 irrespective of the p-value. As the second step, due to a limited sample size, covariates were incorporated into the model one at a time and change in the effect of the exposure on the outcome was measured. If this change was greater than 20% the covariate was left in the model. Since sex of the child and sex of the proband were treated as effect modifiers, we performed stratified analysis of the effect of each androgen on SRS and AOSI: separate models were fit among female children with female probands, female children with male probands, male children with male probands, male children with female probands. In such cases when there were insufficient number of people in a level of a covariate, for example only one person of Hispanic ethnicity among girls with female probands, this covariate was not included in the model to avoid overfitting and inflating standard errors of the model coefficients. In cases where two potential covariates (maternal and gestational age) did not qualify for inclusion into the multiple regression, they were still included to facilitate comparability with earlier published results on the same cohort (27, 40).

### Statistical analyses

Natural log transformation was used to transform the distribution for symmetry of the androgen levels. Both AOSI and SRS was positively skewed, therefore we used natural log transformation of AOSI+1 scores and SRS raw total score, respectively.

Separate robust linear regression models using multi-stage (MM) estimation (41) were fit to assess the association between each of the ln-transformed androgens and ln-transformed AOSI+1 and SRS scores. Since sex of the child and proband’s sex were hypothesized to be effect modifiers, we included a three-way interaction term as well as all lower-level interactions between the variables contained in the three-way term in all the models. In particular, we included four interaction terms in the models: 1) between an androgen and sex of the child, 2) between an androgen and proband’s sex, 3) between sex of the child and proband’s sex, 4) and between an androgen, sex of the child, and proband’s sex. We then assumed that three-way interaction was present if the coefficient of the three-way interaction term was greater than 0 at the significance level of 0.1. Once we found a significant three-way interaction term in several models, the decision was made to fit separate robust regression models within each stratum defined by child’s sex and sex of the proband and present the results within separate strata. Additionally, we fit similar models with untransformed outcomes: total AOSI and total SRS, in order to facilitate more intuitive interpretations. In later sections we discuss the results of the models with untransformed outcomes only if respective results from the models with ln-transformed outcomes were significant at 0.05 level.

Detailed model diagnostics were performed on every model, which included visual and quantitative analysis of residuals, identification of points with high leverage and high influence. Multicollinearity of the multiple regression models was assessed with variance inflation factor using a threshold of 10, and heteroscedasticity was assessed by plotting residuals versus predicted values.

All analyses were performed in SAS version 9.4, and all plots were made in RStudio version 1.0.143. All study participants or their parents/guardians gave their informed consent under the EARLI study approved by the Institutional Review Board of Drexel University.

## Results

### Sample characteristics

Overall and sex-stratified characteristics of the 137 participants (61 females, 76 males) are shown in **Table 1**. Similar to the descriptive results presented by Park *et al*. (42), but using a slightly different analytic sample, there were no sex differences in geometric mean of AOSI score at 12 months: geometric mean (GM)=5.4, geometric standard deviation (GSD)=2.2 among males compared to GM=4.9, GSD=1.9 among females (*P*=0.46). However, boys had on average higher SRS scores than girls: GM=34.4 (GSD=2.0), GM=22.4 (GSD=1.7, *P*<0.0001). Male and female participants had similar distributions of maternal age, gestational age, inter-pregnancy interval, number of pregnancies, proportion of children delivered by cesarean section, maternal race and ethnicity, maternal education, and sex of the proband. However, there were differences in income: girls’ families were more likely to report lower income compared to boys’ families within the middle brackets of income (p=0.03).

**Table 1.** Study characteristics by sex of the child.

| Characteristic | Total |  | Female |  | Male |  | p |
| --- | --- | --- | --- | --- | --- | --- | --- |
|  | Mean | SD | Mean | SD | Mean | SD |  |
| Maternal age | 34.5 | 4.5 | 34.4 | 4.5 | 34.6 | 4.5 | 0.84 |
| Gestational age at delivery | 39.3 | 1.7 | 39.3 | 1.8 | 39.3 | 1.5 | 0.96 |
| Interpregnancy interval | 1671 | 968 | 1737 | 880 | 1617 | 1038 | 0.47 |
| Total number of pregnancies | 3.7 | 1.6 | 3.5 | 1.4 | 3.9 | 1.7 | 0.16 |
| Total AOSI score* | 5.2 | 2.1 | 4.9 | 1.9 | 5.4 | 2.2 | 0.46 |
| Total SRS score* | 28.4 | 2.0 | 22.4 | 1.7 | 34.4 | 2.0 | <0.0001 |
|  | Mean<br>(Median) | SD<br>(IQR) | Mean<br>(Median) | SD<br>(IQR) | Mean<br>(Median) | SD<br>(IQR) |  |
| Total AOSI Score | 5.47<br>(5) | 4.14<br>(3-8) | 4.79<br>(4) | 3.08<br>(3-7) | 6.01<br>(5) | 4.78<br>(2.5-8.5) | 0.33** |
| Total SRS Score | 35.66<br>(28) | 25.75<br>(18-42) | 25.57<br>(22) | 13.15<br>(16-33) | 43.75<br>(35) | 30.24<br>(21-65) | <0.001** |
|  | % | n | % | n | % | n |  |
| Cesarean delivery |  |  |  |  |  |  |  |
| Yes | 35.3 | 48 | 38.3 | 23 | 32.9 | 25 | 0.51 |
| Maternal race |  |  |  |  |  |  |  |
| White | 66.9 | 89 | 61.0 | 36 | 71.6 | 53 | 0.50 |
| Black | 9.8 | 13 | 13.6 | 8 | 6.8 | 5 |  |
| Asian | 12.0 | 16 | 13.6 | 8 | 10.8 | 8 |  |
| Other | 11.3 | 15 | 11.9 | 7 | 10.8 | 8 |  |
| Maternal ethnicity |  |  |  |  |  |  |  |
| Hispanic | 17.5 | 24 | 16.4 | 10 | 18.4 | 14 | 0.76 |
| Maternal Education |  |  |  |  |  |  |  |
| <9th | 3.7 | 5 | 4.9 | 3 | 2.7 | 2 | 0.94 |
| High school graduate | 25.9 | 35 | 26.2 | 16 | 25.7 | 19 |  |
| College | 40.7 | 55 | 39.3 | 24 | 41.9 | 31 |  |
| Graduate/professional degree | 29.6 | 40 | 29.5 | 18 | 29.7 | 22 |  |
| Income |  |  |  |  |  |  |  |
| <\$29,999 | 8.4 | 11 | 6.9 | 4 | 9.6 | 7 | 0.03 |
| \$30,000-49,999 | 16.8 | 22 | 25.9 | 15 | 9.6 | 7 | |
| \$50,000-74,999 | 15.3 | 20 | 15.5 | 9 | 15.1 | 11 | |
| \$75,999-99,999 | 16.0 | 21 | 6.9 | 4 | 23.3 | 17 | |
| \$100,000+ | 43.5 | 57 | 44.8 | 26 | 42.5 | 31 | |
| Sex of the proband |  |  |  |  |  |  |  |
| Female | 16.8 | 23 | 16.4 | 10 | 17.1 | 13 | 0.91 |
| Male | 83.2 | 114 | 83.6 | 51 | 82.9 | 63 |  |

\*Geometric mean and geometric standard deviation
\*\*Two-sided p-value for Mann-Whitney U test

### Meconium androgen measurements

Levels of u-T and u-A4 were positively correlated in the entire sample, as well as among male and female participants (**Table 2**). U-T was positively correlated with u-DHEA among all participants and among girls. Similarly, u-A4 and u-DHEA were positively correlated with each other in the entire sample and among girls. Levels of total androgens were correlated with each other among all children, except for t-A4-t-DHEA. In sex-stratified analysis, total androgens were positively correlated with each other, with the exception of t-T-t-A4 and t-A4-t-DHEA, which were not correlated among girls.

**Table 2.** Spearman rank correlation of unconjugated and total androgens in meconium.

|  | All subjects<br>(n=137) |  |  | Male<br>(n=76) |  |  | Female<br>(n=61) |  |  |
| --- | --- | --- | --- | --- | --- | --- | --- | --- | --- |
|  | T-A4 | A4-DHEA | DHEA-T | T-A4 | A4-DHEA | DHEA-T | T-A4 | A4-DHEA | DHEA-T |
| **Unconjugated** | 0.33** | 0.31** | 0.17* | 0.23* | 0.15 | 0.09 | 0.50** | 0.50** | 0.39* |
| **Total** | 0.27* | 0.16 | 0.45** | 0.40* | 0.26* | 0.47** | 0.18 | 0.02 | 0.45* |

\*p-value < 0.05
\*\*p-value < 0.0001

Both u-T and t-T was higher in males: median u-T=0.57 ng/g (IQR: 0.07-2.27) among boys compared to 0.14 ng/g (IQR: 0.04-1.80) among girls (p=0.01), and median total T=31.97 ng/g (IQR: 10.00-115.67) among boys compared to 13.21 ng/g (IQR: 4.99-66.55) among girls (p=0.02). Levels of meconium u-DHEA were higher for girls (p=0.04): median=10.19 ng/g (IQR: 7.86-17.96), compared to boys: median=8.68 ng/g (IQR: 6.58-12.49). There were no significant differences between u-or t-A4, and t-DHEA between males and females. All of these differences mirror the findings within all participants with meconium samples in EARLI as previously reported. (28)

### Associations of androgen levels and ASD-quantitative phenotype at 12 and 36 months – results of correlation analysis

There was no observed correlation between any of the ln-transformed androgens and 12-month AOSI scores in the full sample, however, after stratification by sex of the younger sibling, a weak negative correlation with t-A4 was seen among females (Spearman *ρ*= −0.32, *P*=0.013) (**Table 1S**). Further stratification by proband’s sex revealed stronger positive, marginally significant correlation with t-DHEA (*ρ*= 0.54, *P*=0.058) among males with female probands. Among females with male probands weak negative correlation with t-A4 (*ρ*= −0.31, *P*=0.03) and weak negative correlation with u-DHEA (*ρ*= −0.32, *P*=0.02) was observed.

Meanwhile, both ln-transformed u-T (**Fig. 1**) and t-T (**Fig. 1S**) were positively associated with SRS in the entire sample (*ρ*= 0.21, *P*=0.02 and *ρ*= 0.18, *P*=0.03, respectively) (**Table 2S**). This relationship persisted among males, but not among females. Further stratification by proband’s sex revealed weak positive correlation among males with male probands (*ρ*= 0.24, *P*=0.06 and *ρ*= 0.26, *P*=0.04 respectively). Negative correlation between ln-transformed t-T and SRS was seen among females with male probands (*ρ* = −0.30, *P*=0.03). Among females with female probands strong positive correlation of SRS with ln-transformed u-T (*ρ*= 0.70, *P*=0.03), t-T (*ρ*= 0.82, *P*=0.004), u-A4 (*ρ*= 0.67, *P*=0.03), and u-DHEA (*ρ*= 0.77, *P*=0.01).

**Figure 1.**
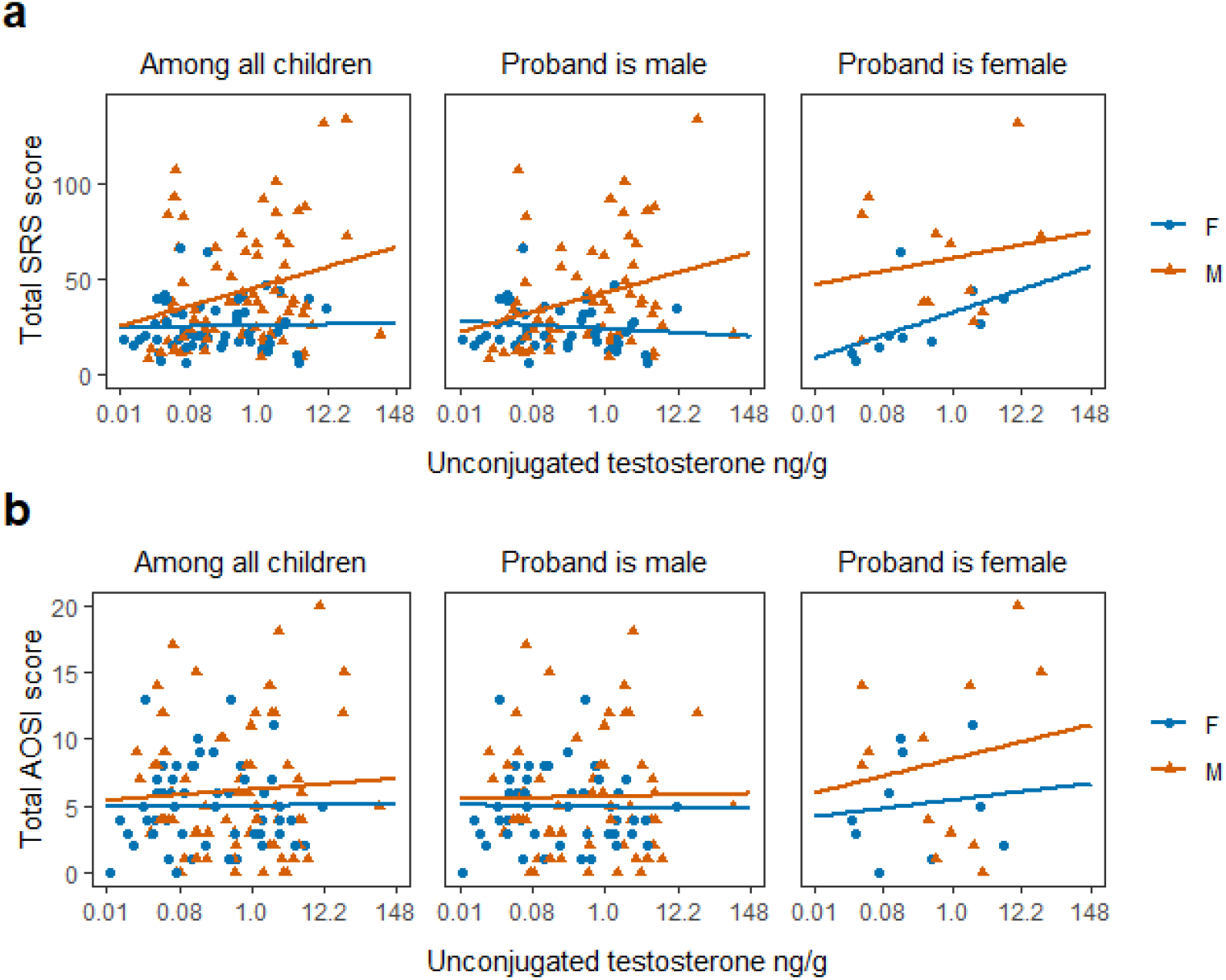
Relationship between meconium unconjugated testosterone (T) and 12- and 36-month outcomes by sex, and further stratified by proband’s sex. **a** Unconjugated testosterone levels vs score on the social responsiveness scale (SRS). **b** Autism Observation Scale for Infants (AOSI).

### Results of unadjusted and adjusted robust linear regression models

The results of robust linear regression models between androgen levels and AOSI score at 12 months are shown in **Figure 2** and **Tables 1S and 2S**. Although no association between androgens and AOSI among females with older affected female siblings was evident in the unadjusted models, positive association was found between total T, unconjugated A4, and unconjugated DHEA and AOSI in this group after adjusting for maternal and gestational age. In fact, the models predicted that with every 25% increase in the levels of total T, unconjugated A4, and unconjugated DHEA, AOSI would increase by 11% (1.25^0.48^ = 1.11, *P* = 0.014), 57% (1.25^2.02^ = 1.57, *P* = 0.01), and 28% (1.25^1.09^ = 1.28, *P*=0.01), respectively. Same adjusted models but with untransformed outcome showed that AOSI will increase by 1.62 (or 0.42 standard deviation of AOSI) with one unit increase in natural logarithm of total T, by 7 (1.82 standard deviations of AOSI) with every unit increase in natural logarithm of unconjugated A4, and by 5.06 (1.32 standard deviations of AOSI) with every unit increase in natural logarithm of unconjugated DHEA (Table 2S). Analogously, in the group of female children with older affected female siblings, the robust regression models revealed positive associations between unconjugated T and SRS in both unadjusted and adjusted analyses (**Figure 3** and **Tables 3S and 4S)**: 6% and 5% increase in SRS with 25% increase in unconjugated T, respectively (1.25^0.26^, *P* < 0.001, and 1.25^0.22^, *P* = 0.004). Similar adjusted model with untransformed SRS showed that SRS will increase by 5.76 points (0.32 standard deviation of SRS) with every unit increase in ln-transformed unconjugated T. In the same group, positive association between total T and SRS was present in the unadjusted model (7% increase in SRS with 25% increase in total T: 1.25^0.31^, *P* = 0.003), but lost statistical significance in the multivariable model (7% increase in SRS with 25% increase in total T: 1.25^0.31^, *P* = 0.10). In the adjusted model with untransformed SRS, one unit increase in natural logarithm of total T predicted 7.3-point increase in SRS (0.41 standard deviations of SRS). Similarly, unconjugated A4 was positively associated with SRS in girls with female probands in the unadjusted analysis predicting 36% increase in SRS with 25% increase in the androgen level (1.25^1.37^ = 1.36, *P* = 0.002), however the relationship was only marginally statistically significant in the adjusted analysis (1.25^1.16^ = 1.30, *P* = 0.07). The adjusted model with total SRS as an outcome predicted that one unit increase in natural logarithm of unconjugated A4 will result in 29.71-point increase in SRS (or 1.67 standard deviations of SRS). In the same group unconjugated DHEA was associated with SRS in both unadjusted and adjusted analyses: the models predicted 23% and 21% increase in SRS with 25% increase in unconjugated DHEA, respectively (1.25^0.92^ = 1.23, *P* = 0.001, and 1.25^0.85^ = 1.21, *P* = 0.007). In the similar adjusted model but with untransformed outcome one unit increase in natural logarithm of unconjugated DHEA predicted 41.95-point increase in SRS (2.35 standard deviations of SRS).

**Figure 2.**
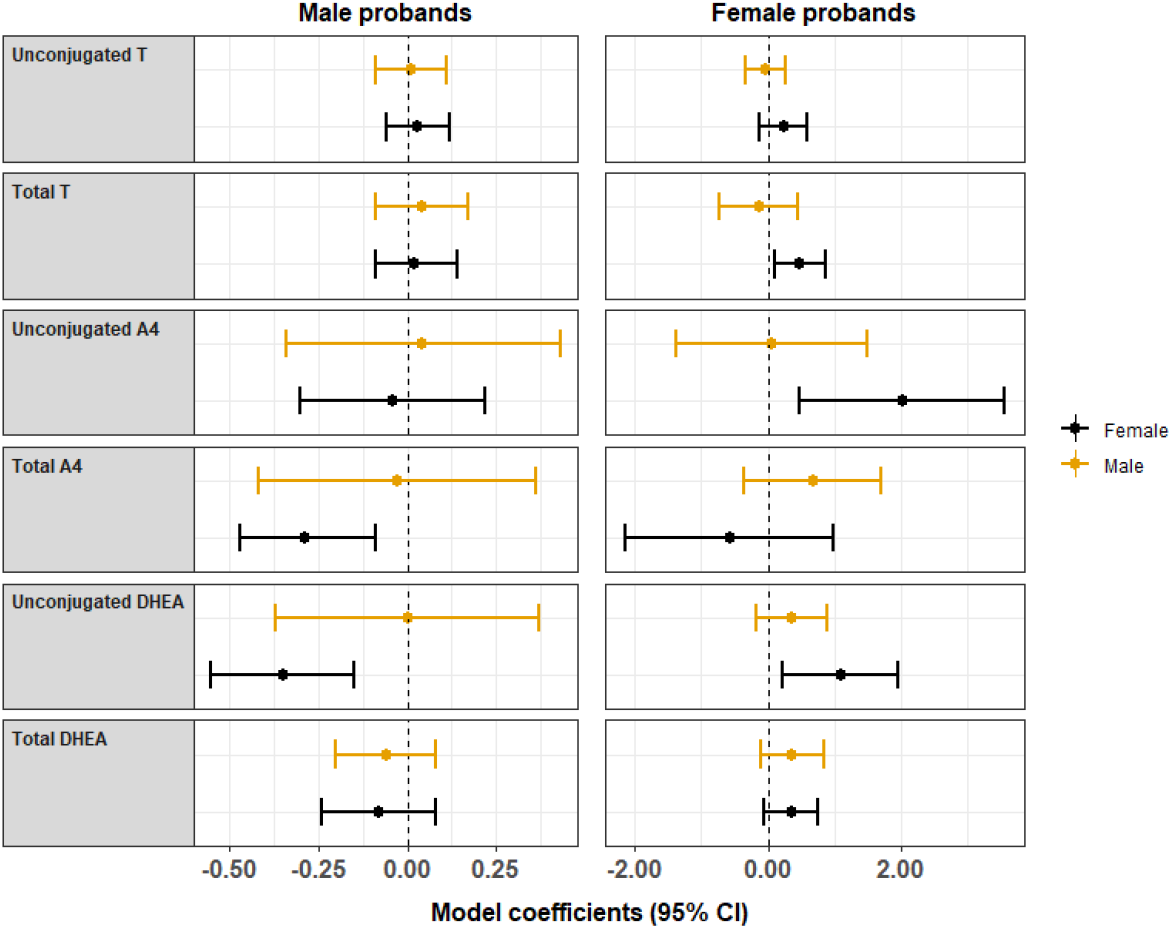
Coefficients and associated 95% Confidence Intervals (CIs) of the ln-transformed androgens from the adjusted robust linear regression model predicting ln-transformed AOSI score. The models were adjusted for maternal and gestational age.

**Figure 3.**
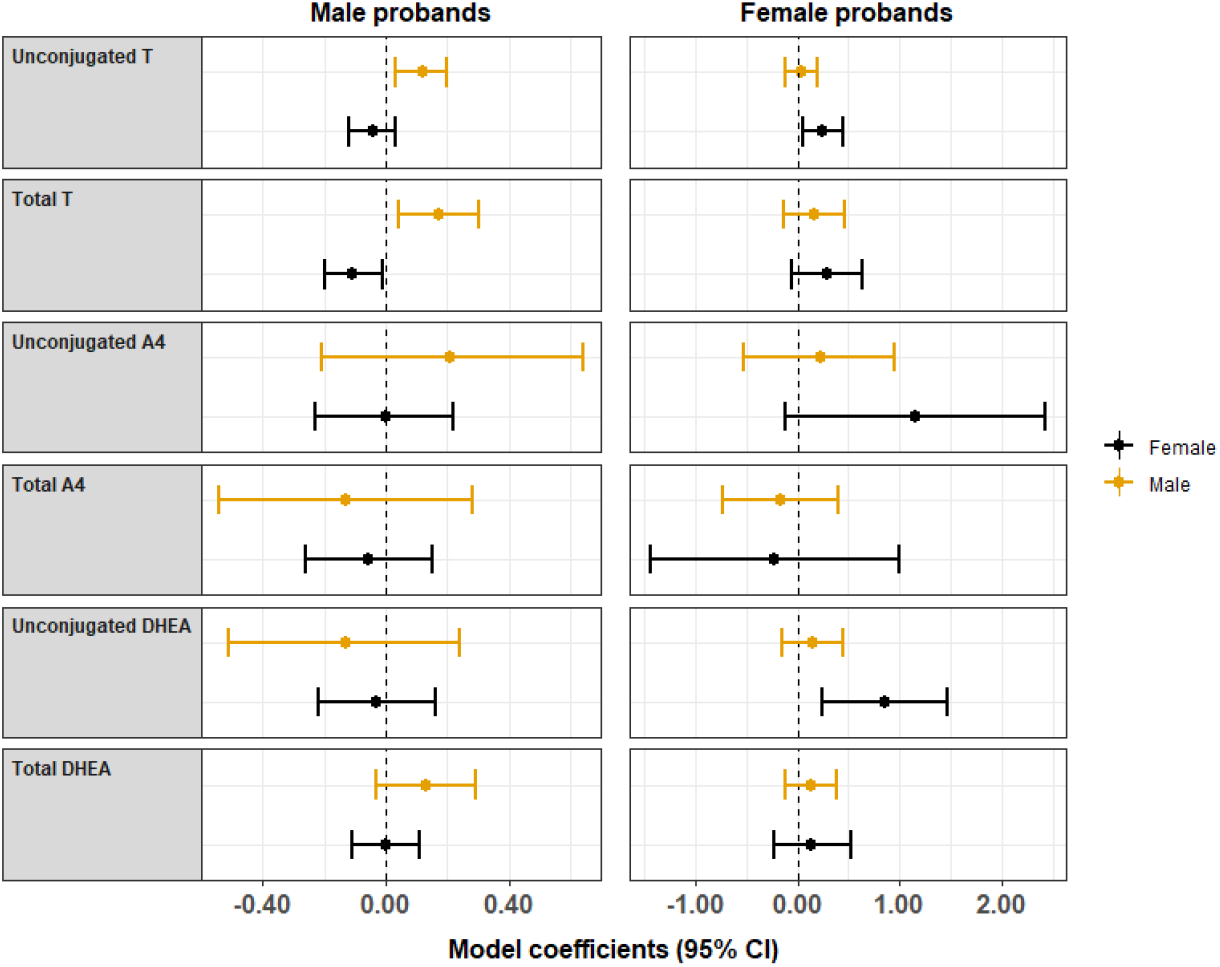
Coefficients and associated 95% Confidence Intervals (CIs) of the ln-transformed androgens from the adjusted robust linear regression models predicting ln-transformed SRS score. The models were adjusted for maternal and gestational age.

In the group of male children with older affected male siblings the regression analyses did not find associations between any of the androgens and the AOSI score, however, unconjugated and total T were associated with SRS in both single and multiple regression models. Both unadjusted and adjusted models predicted 3% increase in SRS score with 25% increase in unconjugated T (1.25^0.12^ = 1.03, *P* = 0.02, and *P* = 0.01, respectively), while 25% increase in total T will result in 3 and 4 % increase in SRS according to the unadjusted and adjusted model, respectively (1.25^0.15^ = 1.03, *P* = 0.04, and 1.25^0.17^ = 1.04, *P* = 0.01). Adjusted models with untransformed SRS showed that one unit increase in natural logarithm of unconjugated T will result in 3.66-point increase in SRS, which translates into 0.13 standard deviation of SRS, while one unit increase in natural logarithm of total T will result in 4.41-point increase in SRS (or 0.15 standard deviation of SRS in this group).

Inverse associations between levels of some androgens and quantitative ASD phenotype were found among females with older affected male siblings. Specifically, AOSI was negatively associated with unconjugated DHEA: the unadjusted model predicted 6% decrease in AOSI with 25% increase in unconjugated DHEA (1.25^*−*0.26^ = 0.94, *P* = 0.01), while adjusted model predicted 8% decrease in AOSI with 25% increase in DHEA (1.25^*−*0.35^ = 0.92, *P* = 0.001). Similar adjusted model but with untransformed AOSI predicted 1.13-point decrease in AOSI (0.38 standard deviation of AOSI) with one unit increase in ln-transformed DHEA. Meanwhile, 25% increase in the levels of total A4 predicted on average decrease of 5% and 6% in AOSI, according to unadjusted and adjusted models respectively (1.25^*−*0.25^ = 0.95, *P* = 0.01, and 1.25^*−*0.29^ = 0.94, *P* = 0.003). According to the adjusted model with total AOSI as an outcome, one unit increase in ln-transformed total A4 will result in 1.19-point decrease in AOSI, which is 0.40 standard deviation of AOSI. Similarly, SRS was negatively associated with total testosterone in this group: both unadjusted and adjusted models predicted an average 2% decrease in SRS with 25% increase in total T (*P* = 0.06, *P* = 0.02, respectively). Meanwhile, adjusted model with untransformed SRS predicted 2.43 decrease in SRS (0.20 standard deviation of SRS) with one unit increase in natural logarithm of total T.

We found no associations between any of the androgen concentrations and quantitative ASD phenotype among males with female probands.

## Discussion

This study investigated the link between meconium androgen concentrations and ASD traits, as measured by AOSI at 12 months and SRS at 36 months, among children whose older sibling was diagnosed with ASD. We built upon previous analysis by including stratification by proband sex and by including both unconjugated and conjugated forms of steroids. These additional analyses recapitulated a previously observed interaction with sex of the proband.

When comparing both the 12 month AOSI scores and the 36 month SRS scores the positive associations were generally observed in either all probands or when the proband was a female, with the one exception being total T measured in male infants having a positive association with SRS scores when the proband was a male. Alternatively, the negative associations we detected occurred when the proband was a male. This trend of positive associations occurring when the proband is a female agrees with a previous study completed with this same EARLI cohort. Park, *et al*., found that the strata with a female proband was observed in a secondary analysis to be the only strata with a detectable association between cord blood unconjugated testosterone and SRS. In meconium from this female proband stratum, associations became apparent and statistically significant that often were not seen in the entire sample or in the strata defined only by sex.

A secondary observation when comparing the 12 month AOSI scores and the 36 month SRS scores is the significance of the infant’s sex. For the AOSI score analysis, both positive and negative associations were generally observed when the infant was a female. However, the SRS scores had significant associations for meconium androgens measured from both male and female infants.

Interestingly, robust linear regression models among female children with female probands consistently estimated high *r*^2^ values even in the crude analysis, indicating that the exposure variable (meconium androgen levels) alone could explain a substantial amount of variability in the ASD-assessment scores in this group. When using AOSI as the outcome, this tendency was observed in the models with total T, unconjugated A4, and total DHEA (the latter association was positive but not significant). When SRS was the outcome, we observed similar phenomenon in the models with unconjugated T, unconjugated A4, and with unconjugated DHEA, in which the association was relatively strong, statistically significant, and with high *r*^2^ in the adjusted and unadjusted analyses. While associations between SRS and unconjugated A4, total T, and total DHEA were not statistically significant in this group, they were positive with high *r*^2^ values.

Importantly, the results of our study may be the link that can potentially reconcile contradictory findings in previous studies. For example, while Auyeung and colleagues consistently found the link between fetal T and ASD-assessment scales in the combined analyses as well as among boys and girls, this relationship could have been driven by the group of boys with male probands, and by the group of girls with female probands. Similarly, while Kung, *et al*.,(26) Whitehouse, *et al*.,(24) and Jamnadass, *et al*.(25) all reported null results in both combined and sex-stratified analyses, the association between fetal T and ASD phenotype may have been masked by the additional interaction with proband sex that was not accounted for. The negative correlation we observed of T and SRS among females with a male proband would be especially problematic in this case.

Effect modification of the association between androgens and ASD-phenotype by sex of the participants was shown in multiple studies,(21, 27, 43-47) including the ones that demonstrated that girls are more susceptible to the effect of abnormal fetal testosterone levels than boys.(21, 43) Effect modification of the association by probands’ sex was reported by Park, *et al*. using the same EARLI study sample as this study,(27) where unconjugated fetal testosterone measured in cord blood was associated with increase in both AOSI and SRS among siblings of ASD-affected females. In addition, there was a marginally significant association between A4 and SRS in the analysis stratified by proband’s sex and adjusted for child’s sex. Thus, within EARLI in both cord blood and meconium, we detect triple interaction between sex steroids, child’s sex, and proband’s sex where the interaction between androgens and sex is different by probands’ sex. The consistency of positive correlation across different androgens, different outcomes in this group, and different biosamples reflecting different windows of prenatal metabolism is worth highlighting. It is now important to replicate these findings using other cohorts, and future studies would benefit from including proband sex as a consideration in study design.

In addition to data scarcity, our study had other limitations. One of them may be the considerable variability of the meconium androgen measurements, even after ln-transformation of the exposure variables. Due to limited sample size, we used robust regression models rather than removal of outliers to alleviate some of these concerns. Since this decision may have a strong effect on our results, we presented full scatterplots of the analyzed data to provide a comprehensive view of the reported relationships. The limited number of studies to date on endogenous metabolites in meconium limits our ability to compare the measures in this cohort versus others. Likewise, the physiological distinction between unconjugated and conjugated steroids in meconium is unclear. Although analyzing both forms of steroids inflated the potential for type I error via multiple comparisons, we thought this was necessitated by the fact that both pools of metabolites were consistently quantifiable within meconium.

Additionally, this study used ASD-assessment scales as outcomes instead of diagnostic categories. While scales, such as AOSI or SRS, provide a continuous measure of ASD-related phenotype, hence increased power for the statistical analysis, the relationship between higher scores on these scales and ASD diagnoses is not unequivocal. However, some studies showed that relationships between certain genetic and environmental exposures and ASD have similar magnitudes irrespective of using ASD diagnoses or continuous scale,(48, 49) suggesting that both higher scores on ASD-assessment scales and ASD diagnosis reflect a similar outcome.

Finally, our findings must be considered within the constraints of the enriched-risk cohort design, meaning that the risk of ASD in this cohort was higher than in the general population by design. If the effect of fetal steroids on ASD phenotype is modified by genetic architecture enriched in this cohort, or severity by selection on the proband, the findings could be attenuated in the broader population of people without family history of ASD. Thus, replication of our findings in studies utilizing samples from a general population may provide evidence supporting generalizability of the results of this study.

## Data Availability

Data is available via National Database for Autism Research (NDAR) and the Steering Committee of EARLI. The corresponding author remains blinded to potentially identifying information.

https://nda.nih.gov/about.html

## Author Contributions

DT and NWS were the lead authors and made substantial contributions to conception, analysis, and interpretation of the data. BKL, BYP, LAC, DMF, IHP, KL and CJN were involved in the conception of the study as well as drafting the manuscript and revising it critically for important intellectual content. AJF made substantial contributions to acquisition and interpretation of data as well as drafting the manuscript. EK, LM, AB, ELV made substantial contributions to study design,acquisition and analysis of data, as well as critically revising the manuscript for important intellectual content. All authors have given the final approval of the version to be published.

## Acknowledgements

Data collection and analyses were supported by NIH R01ES016443, NIH R21ES02559, and Autism Speaks AS5938. BYP was supported by an Autism Speaks predoctoral fellowship (AS 8556). Meconium method development and analysis was supported by R21HD087866, R03CA211820, R01ES029336, and a NARSAD Young Investigator Brain and Behavior Foundation grant to NWS.

## Ethics approval and consent to participate

This study was approved by the Drexel University Institutional Review Board (IRB), and informed consent was obtained from all participants and/or their parent/guardian in accordance with the Drexel University IRB approved protocol.

---

**Figure 1S.**
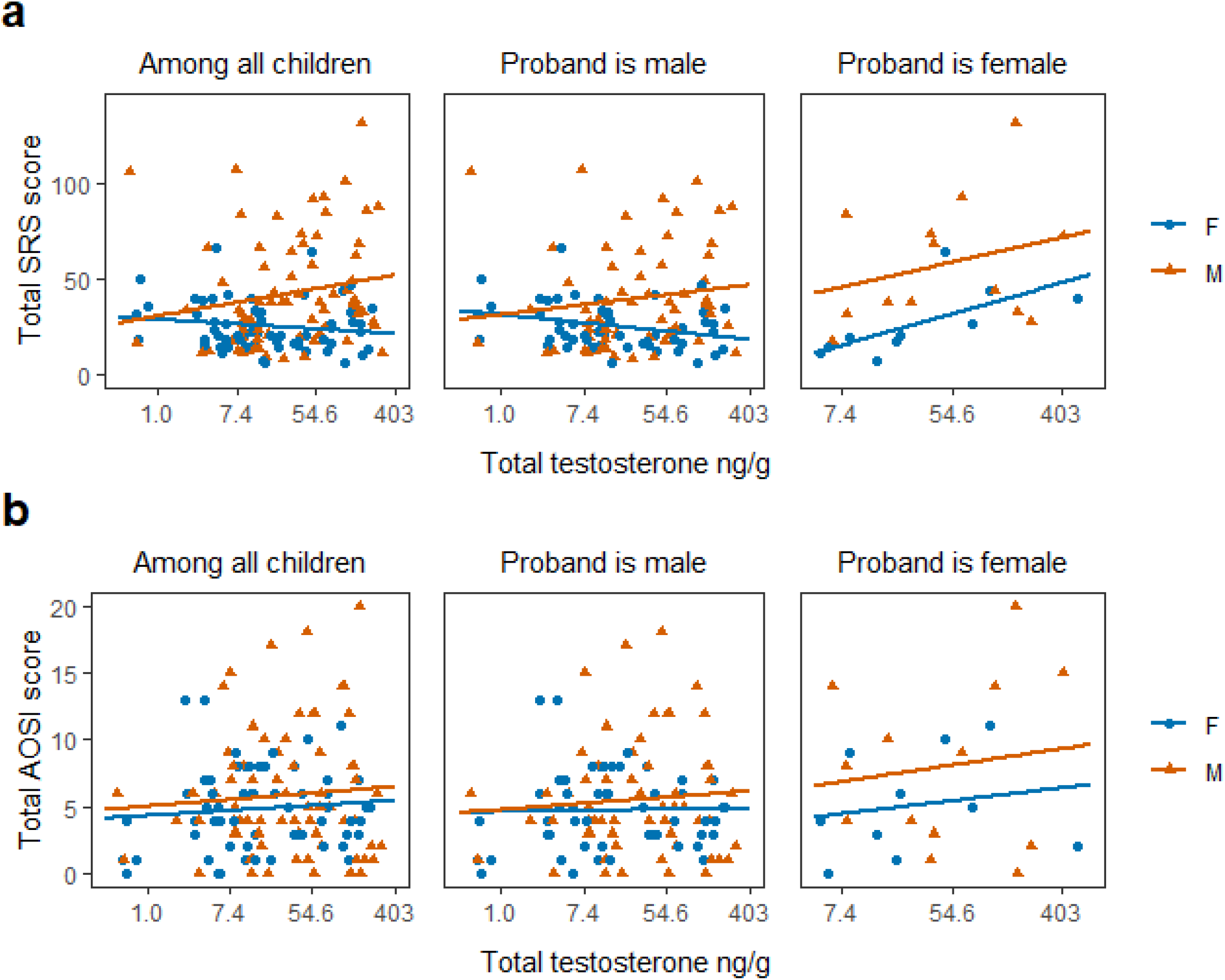
Relationship between meconium total testosterone (T) and 12- and 36-month outcomes by sex, and further stratified by proband’s sex. **a** Total testosterone levels vs score on the social responsiveness scale (SRS). **b** Autism Observation Scale for Infants (AOSI).

**Figure 2S.**
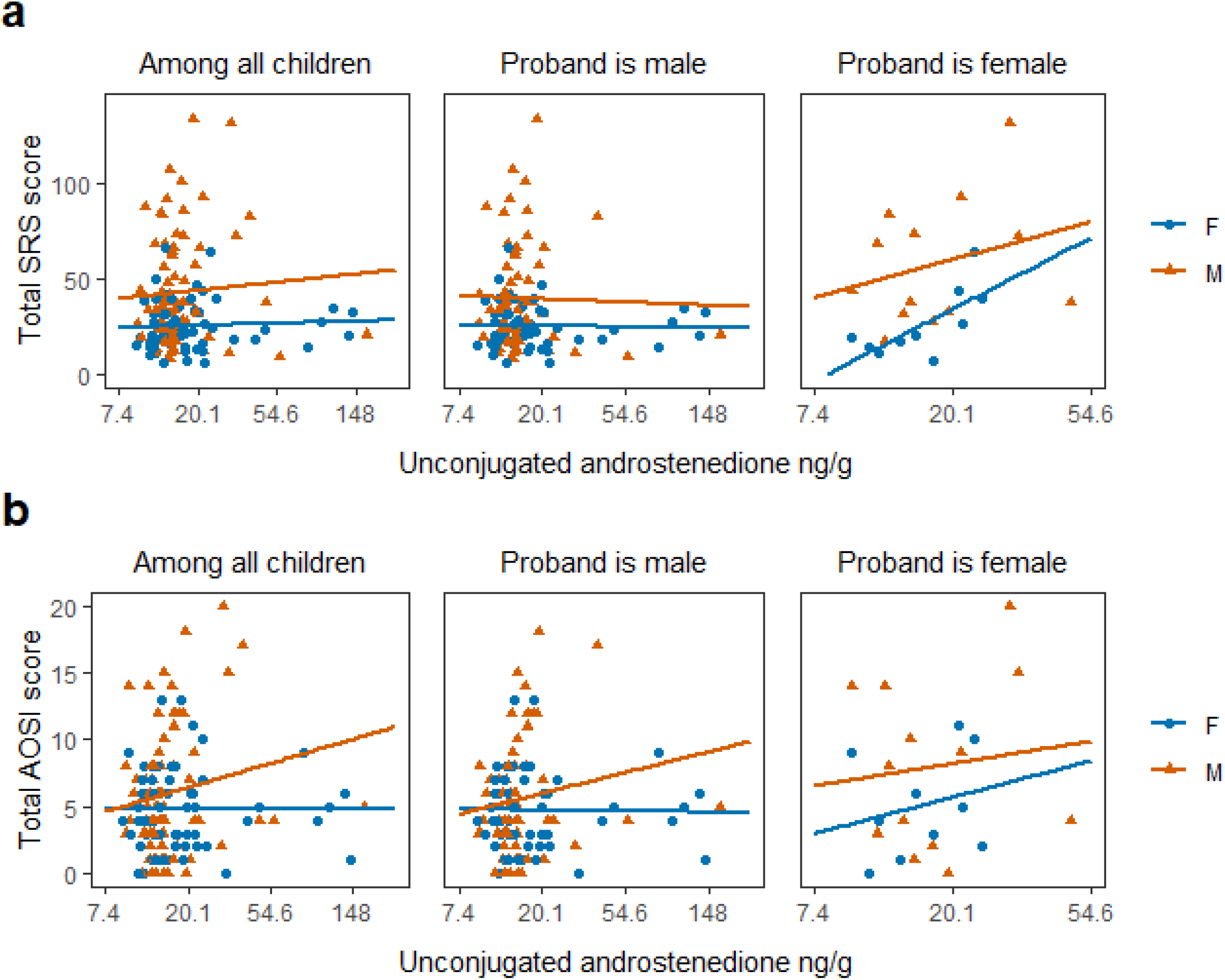
Relationship between meconium unconjugated androstenedione (A4) and 12- and 36-month outcomes by sex, and further stratified by proband’s sex. **a** Unconjugated A4 levels vs score on the social responsiveness scale (SRS). **b** Autism Observation Scale for Infants (AOSI).

**Figure 3S.**
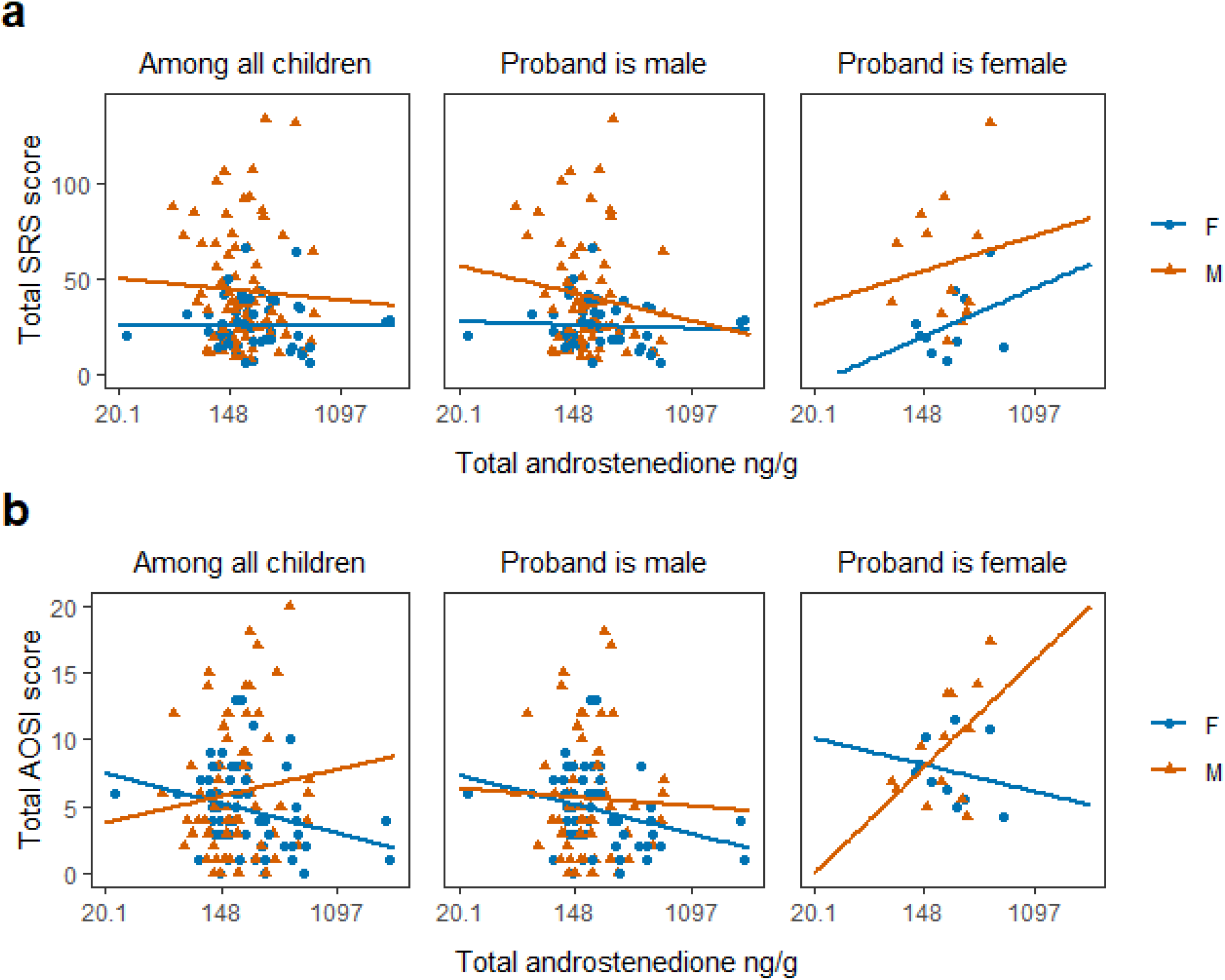
Relationship between meconium total androstenedione (A4) and 12- and 36-month outcomes by sex, and further stratified by proband’s sex. **a** Total A4 levels vs score on the social responsiveness scale (SRS). **b** Autism Observation Scale for Infants (AOSI).

**Figure 4S.**
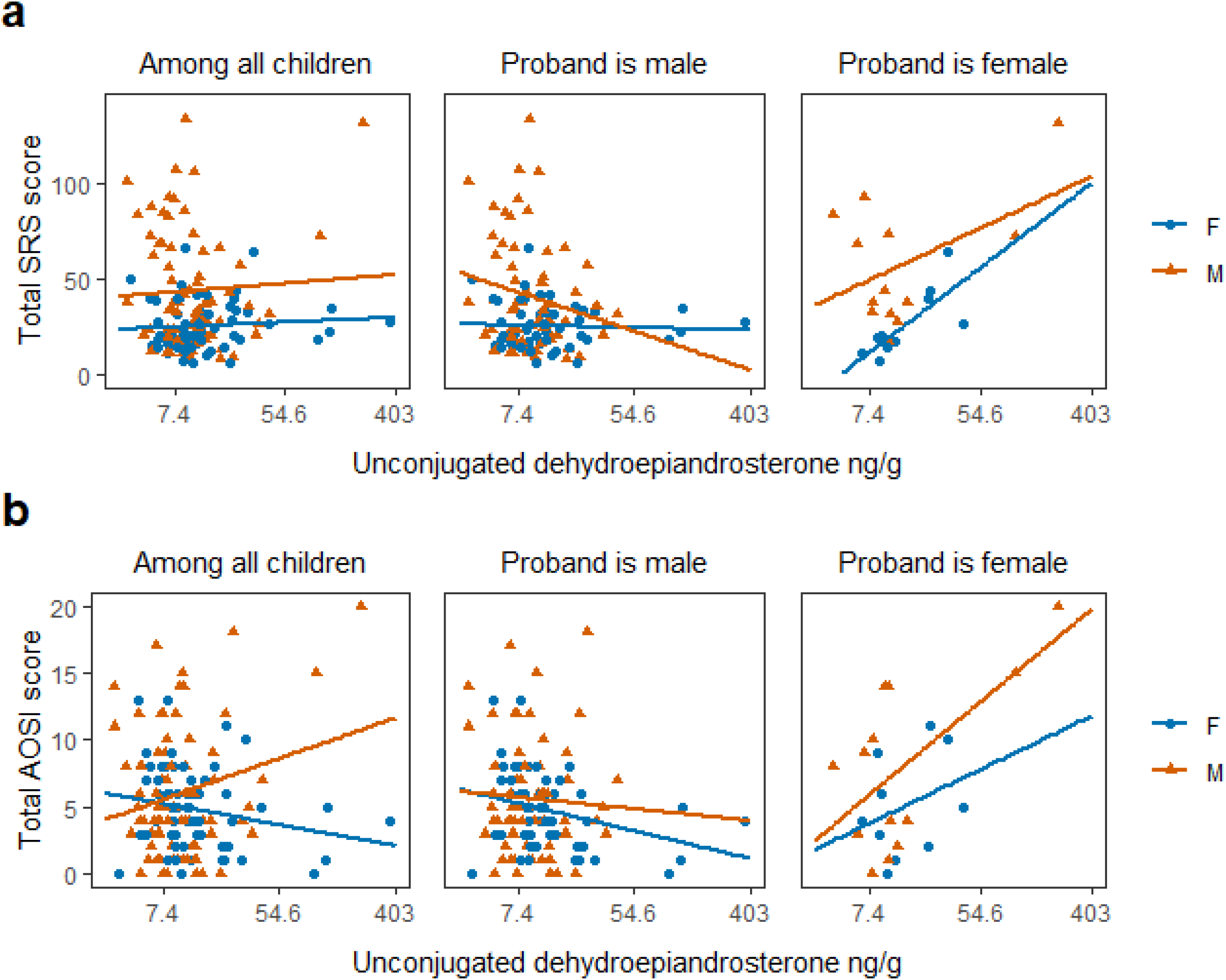
Relationship between meconium unconjugated dehydroepiandrosterone (DHEA) and 12- and 36-month outcomes by sex, and further stratified by proband’s sex. **a** Unconjugated DHEA levels vs score on the social responsiveness scale (SRS). **b** Autism Observation Scale for Infants (AOSI).

**Figure 5S.**
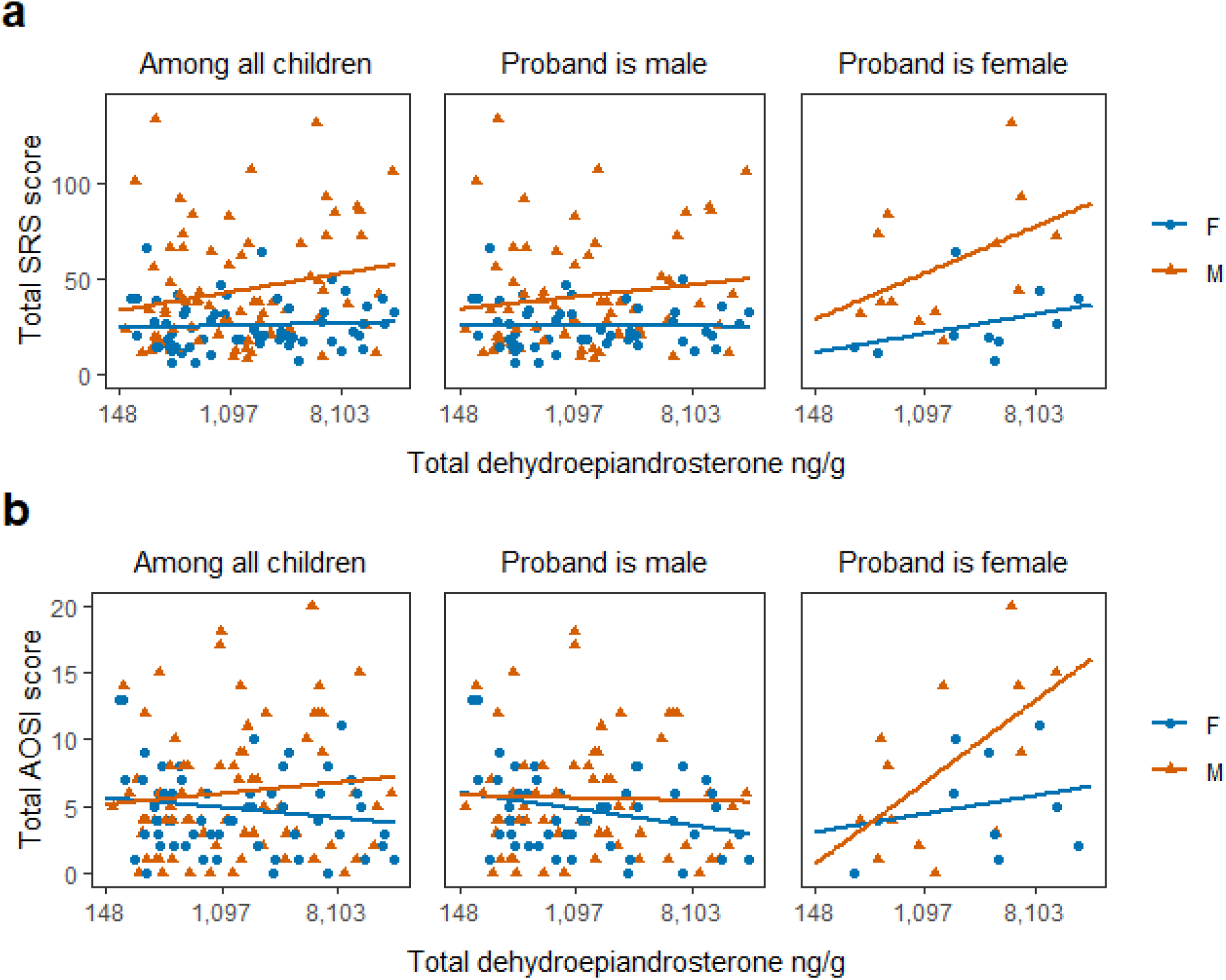
Relationship between meconium total dehydroepiandrosterone (DHEA) and 12- and 36-month outcomes by sex, and further stratified by proband’s sex. **a** Total DHEA levels vs score on the social responsiveness scale (SRS). **b** Autism Observation Scale for Infants (AOSI).

**Table 1S.**
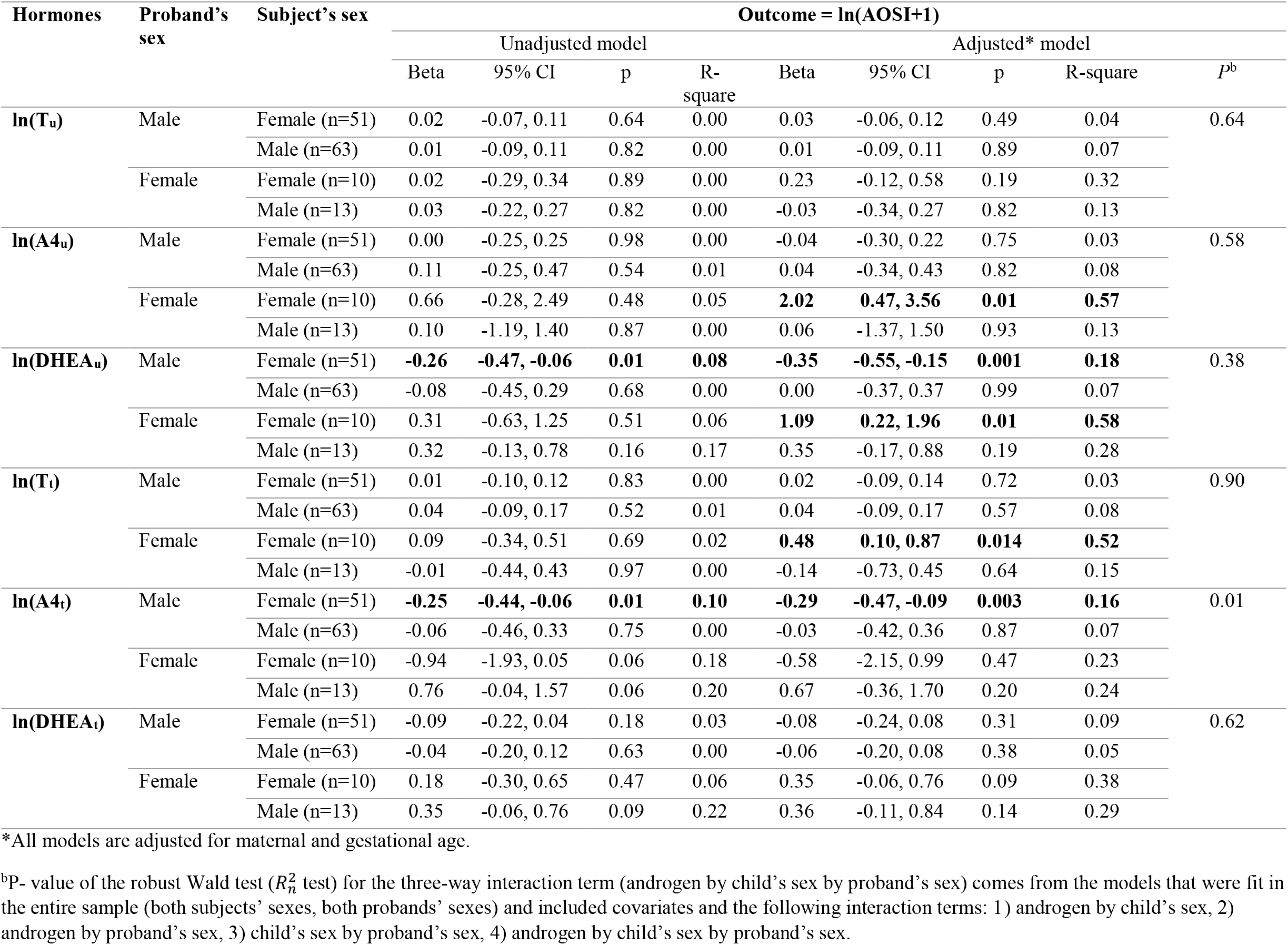
Unadjusted and adjusted log-log models of androgen levels and AOSI score at 12 months by sex of the child and sex of the proband.

**Table 1S.**
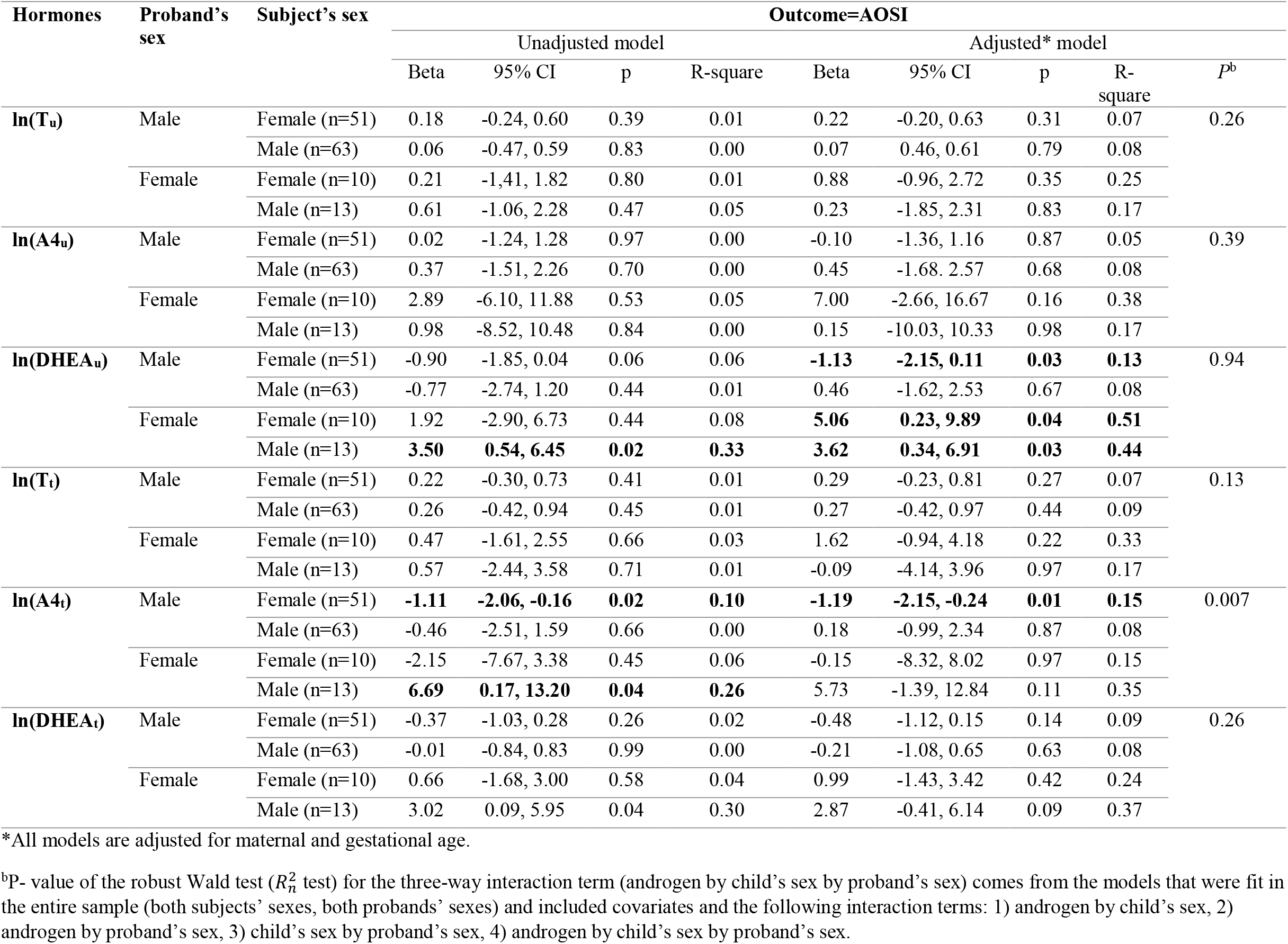
Unadjusted and adjusted models of ln-transformed androgen levels and total AOSI score at 12 months by sex of the child and sex of the proband.

**Table 3S.**
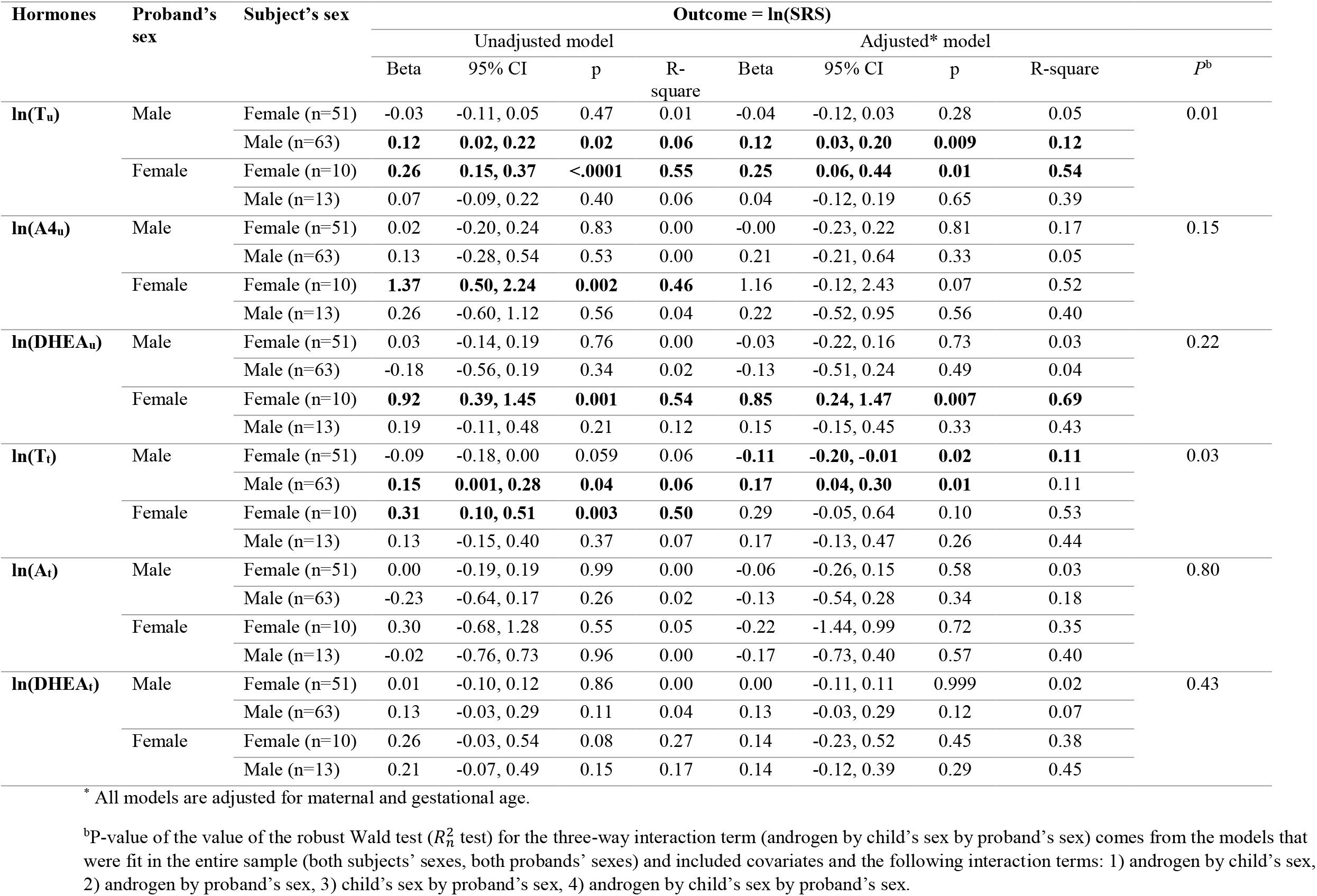
Unadjusted and adjusted log-log models of androgen levels and SRS score at 36 months by sex of the child and sex of the proband.

**Table 4S.** Unadjusted and adjusted models of ln-transformed androgen levels and total SRS score at 36 months by sex of the child and sex of the proband.

| Hormones | Proband's sex | Subject's sex | Outcome = SRS |  |  |  |  |  |  |  | *P*<sup>b</sup> |
| --- | --- | --- | --- | --- | --- | --- | --- | --- | --- | --- | --- |
|  |  |  | Unadjusted model |  |  |  | Adjusted* model |  |  |  |  |
|  |  |  | Beta | 95% CI | p | R-square | Beta | 95% CI | p | R-square |  |
| **ln(T<sub>u</sub>)** | Male | Female (n=51) | -0.60 | -2.35, 1.17 | 0.51 | 0.01 | -0.79 | -2.57, 0.99 | 0.38 | 0.04 | 0.08 |
|  |  | Male (n=63) | 3.27 | -0.08, 6.63 | 0.06 | 0.04 | **3.66** | **0.59, 6.73** | **0.02** | **0.11** |  |
|  | Female | Female (n=10) | **5.48** | **3.34, 7.62** | **<0.0001** | **0.52** | **5.76** | **2.23, 9.30** | **0.001** | **0.46** |  |
|  |  | Male (n=13) | 2.76 | -5.8, 11.33 | 0.53 | 0.03 | 0.02 | -7.86, 7.90 | 0.99 | 0.35 |  |
| **ln(A4<sub>u</sub>)** | Male | Female (n=51) | 0.78 | -4.24, 5.79 | 0.76 | 0.00 | 0.12 | -5.08, 5.32 | 0.96 | 0.02 | 0.17 |
|  |  | Male (n=63) | 4.70 | -9.52, 18.91 | 0.52 | 0.00 | 5.51 | -9.40,20.42 | 0.47 | 0.05 |  |
|  | Female | Female (n=10) | **33.62** | **6.56, 60.68** | **0.01** | **0.37** | 29.71 | -6.88, 66.31 | 0.11 | 0.46 |  |
|  |  | Male (n=13) | 13.15 | -32.87, 59.17 | 0.58 | 0.03 | 7.96 | -29.50, 45.42 | 0.68 | 0.36 |  |
| **ln(DHEA<sub>u</sub>)** | Male | Female (n=51) | 0.68 | -3.16, 4.51 | 0.73 | 0.00 | -0.79 | -5.25, 3.68 | 0.73 | 0.03 | 0.001 |
|  |  | Male (n=63) | -6.78 | -18.77, 5.21 | 0.27 | 0.02 | -2.69 | -14.50, 9.11 | 0.65 | 0.05 |  |
|  | Female | Female (n=10) | **33.45** | **23.70,43.20** | **<0.0001** | **0.47** | **41.95** | **29.76, 54.13** | **<0.0001** | **0.53** |  |
|  |  | Male (n=13) | 13.75 | -3.12, 30.63 | 0.11 | 0.16 | -7.24 | -25.50, 11.01 | 0.44 | 0.35 |  |
| **ln(T<sub>t</sub>)** | Male | Female (n=51) | -2.09 | -4.19, 0.03 | 0.051 | 0.06 | -2.43 | -4.56, -0.29 | 0.26 | 0.10 | 0.011 |
|  |  | Male (n=63) | 4.37 | -0.15, 8.9 | 0.058 | 0.04 | **4.41** | **0.24, 8.58** | **0.04** | **0.10** |  |
|  | Female | Female (n=10) | **6.90** | **3.90, 9.90** | **<0.0001** | **0.49** | 7.3 | -0.11, 14.71 | 0.053 | 0.45 |  |
|  |  | Male (n=13) | 5.11 | -9.68, 19.90 | 0.50 | 0.04 | 4.88 | -9.80, 19.55 | 0.51 | 0.37 |  |
| **ln(A<sub>t</sub>)** | Male | Female (n=51) | -0.05 | -4.38, 4.28 | 0.98 | 0.00 | -1.27 | -5.88, 3.34 | 0.59 | 0.03 | 0.25 |
|  |  | Male (n=63) | -9.67 | -22.63, 3.29 | 0.14 | 0.02 | -4.41 | -17.52, 8.69 | 0.51 | 0.05 |  |
|  | Female | Female (n=10) | 10.91 | -14.78, 36.61 | 0.41 | 0.07 | -1.72 | -34.49, 31.06 | 0.92 | 0.28 |  |
|  |  | Male (n=13) | -2.34 | -41.64, 36.95 | 0.91 | 0.00 | -16.35 | -36.99, 4.28 | 0.12 | 0.43 |  |
| **ln(DHEA<sub>t</sub>)** | Male | Female (n=51) | 0.57 | -2.02, 3.15 | 0.67 | 0.00 | 0.31 | -2.32, 2.94 | 0.82 | 0.02 | 0.93 |
|  |  | Male (n=63) | 4.94 | -0.41, 10.28 | 0.07 | 0.03 | 4.29 | -0.98, 9.56 | 0.11 | 0.07 |  |
|  | Female | Female (n=10) | **6.07** | **0.09, 12.04** | **0.047** | **0.25** | 3.36 | -6.15, 12.88 | 0.49 | 0.31 |  |
|  |  | Male (n=13) | 10.81 | -4.64, 26.27 | 0.17 | 0.15 | 6.08 | -7.95, 20.11 | 0.40 | 0.39 |  |

\* All models are adjusted for maternal and gestational age.
<sup>b</sup>P-value of the robust Wald test ( $R_n^2$ test) for the three-way interaction term (androgen by child's sex by proband's sex) comes from the models that were fit in the entire sample (both subjects' sexes, both probands' sexes) and included covariates and the following interaction terms: 1) androgen by child's sex, 2) androgen by proband's sex, 3) child's sex by proband's sex, 4) androgen by child's sex by proband's sex.

## Notes

### Competing Interest Statement

The authors have declared no competing interest.

